# Varicella outbreak at a nursery school under routine immunization

**DOI:** 10.1101/2021.05.07.21256754

**Authors:** Tomoko Sakaue, Tamie Sugawara, Yoshiyuki Sugisita, Junko Kurita, Michiko Nohara, Yasushi Ohkusa

## Abstract

**Background and objective:** In Japan, routine administration of two-dose immunization for varicella to one-year-old children was introduced in October, 2014. The objective of this study was measurement of the effectiveness of routine two-dose immunization for varicella to onset and assessment of severity in a nursery school setting.

**Method:** The study period extended from the beginning of April, 2017 through March, 2018. The study area was Nursery school B in a city A. Subjects were 120 children in all. We analyzed vaccine efficacy (VE) as an observational study and assessed severity using Fisher’s exact test. We also assessed VE for severity using linear regression. Severity was defined as the length of school absence attributable to varicella infection.

**Results:** For one dose or more, VE was 48.1% for all ages and 49.2% among children three years old and older. No significant VE was found. Vaccination using one dose or more can reduce severity significantly.

**Discussion and conclusion:** Low VE was found in a nursery school setting, although results were not significant. VE for severity was confirmed, but a second dose might not reduce severity.

## Introduction

Before the advent of routine immunization, varicella was a commonly encountered pediatric infectious disease caused by varicella-zoster virus [1]. A policy of routine two-dose immunization for varicella to one-year-old children was put into effect in Japan in October, 2014. Before routine immunization, varicella outbreaks occurred every year. At that time, national official sentinel surveillance reported 0.2 million patients at a sentinel medical facility. After initiation of the routine immunization policy, cases fell to approximately one-third of the earlier frequency. Moreover, the age distribution of patients changed. Before initiation until 2014, children younger than four years old accounted for about 85% of all cases. After initiation, in 2017, their share was about 40%.

Moreover, number of 5–14-year-old patients decreased, but their proportion increased. Other countries that introduced routine immunization for varicella reported similar outcomes [2]. Although routine immunization greatly reduces varicella prevalence, the booster effect declines. Cost-effectiveness analyses including specific assessments of booster effects and herpes zoster were also conducted [3].

In 2018, when several years had passed after initiation of routine immunizations, Nursery school B in city A experienced a large outbreak of varicella. As one might expect, a high proportion of children in B had already received the vaccine twice. Fortunately, (nursery) schools in city A activated daily surveillance using the (Nursery) School Absenteeism Surveillance System ((N)SASSy) recording the numbers of patients at (nursery) schools with infectious diseases and presenting information in real time.

The objective of this study was measurement of the effectiveness of two-dose routine immunization for varicella in terms of onset and severity in the nursery school setting using information recorded at (N)SASSy.

## Method

We conducted an observational study of a nursery school during a period extending from the beginning of April 2017 through March 2018 using information from (N)SASSy and health and immunization records. The outbreak circumstances in city A were ascertained through (N)SASSy. The definition of varicella infection was based on the doctors’ clinical diagnoses. Study subjects were children younger than 15 years old. Children five years old and younger attend nursery schools. Children of 6–12 years old attend elementary school. Children who are 13–15 years old attend junior high school.

The outbreak circumstances at Nursery school B were characterized by nursery school health records. The length of absence attributable to varicella was recorded in the health record. Confirmation of recovery and attendance at the nursery school again was done by a doctor. We used the length of absence attributable to varicella as an indicator of varicella symptom severity. Because absences might include weekends, we adopted two measures: one limited to weekdays and one including weekends and holidays. The circumstances of vaccination of varicella at Nursery school B were also provided from immunization records based on the maternity passbook. The dispositions of data from children who were infected before the study period and children who were vaccinated during the study period are explained below.

We assessed vaccine efficacy (VE) based on data from health records and immunization records at Nursery school B using Fisher’s exact test. VE is defined as one minus the incidence among vaccinated children divided by the incidence among unvaccinated children. Incidence was defined as the proportion of infected children in the study period among the population, excluding those who were infected before the study period.

We also calculated VE for one dose only or two doses only, as well as one dose or more. When the vaccination was limited to “one dose only”, the children vaccinated with two doses were excluded from analyses. Similarly, when the vaccinated children were limited to “two doses only”, the children vaccinated with only one dose were excluded from analyses.

We also analyzed VE for severity using linear regression. VE for severity was defined as one minus the incidence with severe symptoms among vaccinated children divided by the incidence with severe symptoms among unvaccinated children. Severity was defined according to the length of absence. However, because results might be affected by the day of the week or by national holidays, we examined two versions of length of absence, respectively including and excluding weekends and holidays. We also examined specifications of four types for each type of severity definition: 1) univariate regression with a dummy variable for one dose or more of vaccine received, 2) univariate regression with a dummy variable for two-dose vaccination received, 3) multivariate regression with a dummy variable for one dose or more of vaccine received and a dummy variable for two-dose vaccination received, and 4) multiivariate regression with a dummy variable for one dose or more of vaccine received and a dummy variable for children three years old and older. Reference categories were “unvaccinated” in specifications 1) – 3) and “unvaccinated younger than three years old” in specification 4). All specifications were estimated using ordinary least squares method. We adopted 5% as the level for inference of significance.

## Ethical considerations

Nursery schools in Japan must manage children’s health condition and immunization records as a measure for infectious diseases [4]. All information about absences, infected diseases, and immunization were anonymized for analyses to prevent use of personal information. Data in (N)SASSy include no personal information about students or nursery school children. This study was approved by the ethical committee of Tokyo Kasei University on February 21, 2020 (No.2019-33).

## Results

For the study period, there were 7,058 0–5-year-old children, 22,323 6–12-year-old children, and 10,355 13–15-year-old children in city A. Nursery school B had 9 children of age 0, 16 children of age 1, 20 children of age 2, 22 children of age 3, 25 children of age 4, and 28 children of age 5. Thirteen children of Nursery school B had been infected with varicella before the study period.

Figure 1 depicts the epidemic curve for Nursery school B. The initial case in the study period was that of a 6-year-old child on 23 December, 2017. A 5-year-old child had onset at three days after the initial case’s onset. Then a 2-year-old child reported onset the following day. A 4-year-old child reported onset the following day. Subsequently, children reported onset at about two week intervals. The first peaks were recorded on about 10 January, consisting mainly of 4–5-year-old children. The second peak appeared on about 22 January, consisting mainly of 3–4-year-old children. Then the number of patients decreased after the second peak. Infections ceased on 18 February because no new patient showed onset during the four weeks prior, which is twice the average length of the incubation period of varicella. Incidence of varicella at Nursery school B during the study period was 0% for age-0 children, 6.25% for age-1, 20.0% for age-2, 36.4% for age-3, 52.0% for age-4, and 42.9% for age-5 children. In all, 38 children were infected during the study period. The age of highest prevalence was four years old, followed by five years old.

**Figure 1:** Epidemic curves by age in nursery school B and in city A from 28 days before onset of the initial cases in Nursery school B. Note: Bars represent the epidemic curve in nursery school B by age: black bar indicates five years old, dark gray bar indicates four years old, light gray bar indicates three years old, bar with vertical lines indicates two years old and bar with transverse lines indicates one year old. Dots represent the epidemic curve in city A from 28 days before the initial cases in nursery school B onset. Especially, the dot shape signifies the facility type: circles represent children in other nursery schools, triangles represent elementary school students and squares represent junior high school students.

Figure 2 shows the varicella incidence in city A by age during the study period. The incidence in 5-year-old children, 2.47%, was highest, followed by 2.02% for 4-year-old children, and 1.81% for 3-year-old children. The incidence in 0-year-old children was 1.69%. Among students, the highest incidence was shown for the second grade of elementary school: 1.72%. That finding also indicates that the outbreak in Nursery school B was larger than the community outbreak for the whole of city A because incidences under five years old in city A were lower than in Nursery school B.

**Figure 2:** Incidence rate by age in nursery school B and in city A. Note: Bars represent the incidence rate in nursery school B. Lines represent the incidence rate in city A. From April 2017 through March 2018, the scale for the incidence rate at nursery school B is shown on the left-hand side. The scale for the incidence rate in city A is shown at the right-hand side.

Figure 3 portrays the epidemic curve for city A. It shows the outbreak from the last half of April–May. It occurred again during December–January.

**Figure 3:** Epidemic curves in city A by age group. Note: Age groups were defined as nursery school children, elementary school students, and junior high school students. Black bars indicate the number of patients at elementary school, dark gray bars indicates the number of patients at junior high school, and light gray bars indicates the number of patients at nursery school.

At Nursery school B, no case arose during one month to two weeks before the initial case had onset in the nursery school on 23 December. During the same period, 16 cases were identified at other nursery schools, 45 cases at elementary schools, and one case at the junior high school.

Length of absence because of varicella infection at Nursery school B, excluding weekends and holidays was 3.92 days on average. The minimum was one day. The maximum was seven days. If including weekends and holidays, it was 5.86 days on average, with the minimum of three days and the maximum of nine days.

Nursery school B retained immunization records for varicella for 107 children. Of those, 104 children (97.2%) received one dose or more of varicella vaccine. Three children did not receive it. The coverage rates by age were 100% among children of age 0, 93.8% for age 1, 94.8% for age 2, 95% for age 3, 100% for age 4, and 95.2% for age 5. Thirteen children who had been infected before the study period were excluded from the analysis presented below. No child was vaccinated during the study period.

Table 1 presents the estimated VEs and its 95% confidence intervals. For one dose or more, it was 48.1%. If limiting the vaccinated children to those who received only one dose, then it was 62.5%. However, if limiting the vaccinated children to those who received the vaccine twice, then results were 42.7%. No estimated VE was found to be significant.

**Table 1:** Vaccine efficacy by number of doses of vaccine in Nursery school B

| Definition of vaccinated | Vaccine efficacy (%) | *p*-value | 95% CI |  |
| --- | --- | --- | --- | --- |
|  |  |  | Lower | Upper |
|  |  |  | bound | bound |
| One dose or more | 48.1 | 0.287 | -0.206 | 0.777 |
| One dose only | 62.5 | 0.195 | -0.046 | 0.866 |
| Two doses only | 42.7 | 0.558 | -0.339 | 0.756 |

Note: “*p*-value” represents a *p*-value obtained from Fisher’s exact test. When the vaccination was limited to “one dose only” at the second row, the children vaccinated with two-dose vaccination were excluded from analyses. Similarly, when the vaccinated children were limited to “two doses only” at the bottom row, the children vaccinated with only one dose were excluded from analyses.

Table 2 presents the estimation results of severity, which was inferred from the length of absence, by definition of severity, and by definition of the vaccinated. Results indicate that children vaccinated with one dose or more had a more than two-day-shorter period of illness than unvaccinated children, both when including and excluding weekends and holidays. Alternatively, if limiting the analyses to those who received two doses of vaccine, then vaccination did not significantly affect severity. When we examined both vaccination classifications at the same time, only the one dose or more of vaccine classification was found to have significant effects by the definition of severity excluding weekends and holidays. However, one dose or more was marginally significant given the definition of severity including weekends and holidays.

**Table 2:** Estimation results for vaccination and severity

| Explanatory variable | Estimated coefficient | *p*-value | 95% CI |  |
| --- | --- | --- | --- | --- |
|  |  |  | Lower | Upper |
|  |  |  | bound | bound |
| Dependent variable: Length of absence excluding weekends and holidays |  |  |  |  |
| One dose or more | -2.194 | 0.016 | -3.959 | -0.429 |
| Two doses | -0.685 | 0.164 | -1.664 | 0.292 |
| One dose or more | -2.000 | 0.047 | -3.970 | -0.029 |
| Two doses | -0.241 | 0.629 | -1.276 | 0.793 |
| One dose or more | -2.111 | 0.014 | -3.760 | -0.462 |
| Three years old or older | 0.427 | 0.016 | 0.083 | 0.770 |
| Dependent variable: Length of absence including weekends and holidays |  |  |  |  |
| One dose or more | -2.250 | 0.015 | -4.033 | -0.466 |
| Two doses | -0.900 | 0.068 | -1.872 | 0.071 |
| One dose or more | -1.857 | 0.064 | -3.829 | 0.115 |
| Two doses | -0.488 | 0.346 | -1.523 | 0.548 |
| One dose or more | -1.857 | 0.064 | -3.998 | -0.428 |
| Three years old or older | -0.4898 | 0.346 | -0.182 | 0.560 |

Note: Dependent variables of the upper panel were the length of absence excluding weekends and holidays. Those of the lower panel were length of absence including weekends and holidays.
Reference categories were “unvaccinated” in the first to third specifications in both panels and “unvaccinated younger than three years old” in the bottom specification in both panels. All specifications were estimated using ordinary least squares method.

## Discussion

Results of this study revealed an association between a community outbreak and an outbreak at a nursery school. The initial case in the nursery school was not found to have been caused by exposure to children in the nursery school. During the one month preceding a period of two weeks before the initial case at the nursery school, there were 16 cases in nursery schools, 45 cases in elementary schools, and one case in junior high schools in city A. One of them might have contacted the initial case or followed a few cases during the two weeks preceding the initial discovery of a case. Subsequently, the following cases had onset without a two-week interval after the initial case onset.

Although VE for infection was around 50%, all estimated VE were not significant. Especially, VE of two-dose vaccination only was lower than that of one-dose vaccination only, although the differences were not significant because these 95%CIs were overlapped. Formally, when we logistically regress onset on dummy variables for one dose only or two doses only, the *p*-value for which the null hypothesis that the two estimated coefficients are the same is 0.2146.

However, higher vaccine efficacy of two doses than of one dose has been reported [5,6]. One report described 94.9% for a second dose vs. 65.4% for one dose for all varicella and 99.5% vs. 90.7% for moderate or severe varicella. Differences in VE between the numbers of doses were significant. The latter were reported as 95.0% vs. 67.0% for all varicella and 99.0% vs. 90.3% for moderate or severe varicella. The difference in VE was significant.

Why have these counter-intuitive phenomena been found from this study? The findings might be attributable mainly to the small sample. If one examines a larger sample in a similar setting, then this phenomenon might be resolved. Alternatively, it simply represents a difference in age in both categories. Unfortunately, when adding a dummy variable for any age or older to the logistic regression described above, this phenomenon cannot be resolved. Another possibility for a different VE pattern among the number of doses might be the type of vaccine. A marked point of difference between the two earlier studies and the present study is that the earlier studies used measles–mumps–rubella–varicella (MMRV) vaccine, but mono-valent vaccine was used for the latter. Regarding the study design, randomized control trial vs. observational study might affect differences in results obtained for VEs. Investigation of this counter-intuitive phenomenon remains as a subject for future research.

Severity was reduced considerably by one dose or more of vaccine. In fact, children who received one dose or more of vaccine were ill for two days fewer than unvaccinated children were. Nevertheless, two-dose vaccination did not have a significant additional effect beyond that of one-dose vaccination.

We defined severity as the length of absence. Because of restrictions imposed by the School Health and Safety Act, varicella-infected students cannot attend school until a rash is crusted. Although this act does not apply to nursery schools, the guidelines for infection control at nursery schools [4] require a similar criterion for nursery school children. Especially for working caregivers, the length of absence of a varicella-infected child reflects the length of absence from a caregiver’s workplace. In this sense, length of absence is a proxy for the amount of rash and the disease burden. Therefore, it was presumed to be an appropriate measure for severity. Nevertheless, if we were to use some information about the number of rashes, unlike the present study [7,8], then the number of rashes would be a more appropriate measure of severity.

At Nursery school B, although the coverage was quite high, many children were infected. Therefore, the vaccination effectiveness was apparently low. However, when considering symptom severity, one-dose vaccination at least reduces severity. Secondary vaccination showed no effectiveness. Because the varicella vaccine used in Japan is an attenuated live vaccine, the antibody positive rate with one-dose vaccination was higher than 90%, but half of the vaccinated children were infected at nursery schools [9]. Even if they had onset, their symptoms were mostly mild, with fewer than 50 rashes. The absence of each was shorter than four days. At Nursery school B, because the average length of absence was 3.92 days, many cases were presumed to be mild. At three preschools in Turkey, VE for one-dose vaccination was 33.6% against varicella disease of any severity and 82.5% against moderate or severe varicella [10]. Our point estimation of the one-dose or two-dose vaccination was comparable to that reported from an earlier study for one-dose vaccination at another nursery school [9], although our estimated VE was not significant. This result might indicate that two-dose vaccination does not raise effectiveness, at least in a nursery school setting.

A case-control study in Japan [11] indicated VE as 76.9% for one-dose vaccination and 94.7% for two-dose vaccination. By contrast, our obtained results indicate that vaccine effectiveness might be much lower, although the results were not found to be significant. This difference might reflect whether the infected children attend nursery school or not. If that case-control study included children who did not attend nursery school, then the frequencies of contact with other children were much different among children who attended nursery school and those who did not. In general, a case-control study is more reliable than an observational study, such as the present study. However, if a case-control study were to include children who did not attend nursery school, then the difference in study settings might greatly affect the results.

In the US, routine immunization for varicella was begun in 1996. In 2000, although the coverage was 80%, the number of patients was reduced by 71–84% [12,13]. Moreover, the number of hospitalizations, fulminant cases, and fatality cases decreased. Therefore, the vaccine effectiveness for avoiding severe illness is apparently large. Similarly, we can confirm VE to reduce illness severity even if the definition of severity was much different in the study conducted in the US.

We used information from (N)SASSy, which is useful for teachers and the persons concerned, such as officers at public health centers and local governments, to prevent the spread of infectious diseases [14–16]. Demonstrably, (N)SASSy is useful as a form of syndromic surveillance in Japan. In short, all information related to children’s health conditions is integrated at (nursery) schools. The system can be a powerful public health tool for use during mass gatherings or important political events [17] such as the G7 summit meeting. In fact, (N)SASSy was developed by a research group headed by Dr. Ohkusa, one author of this paper. It has been funded by the MHLW since 2007, although its copyright has been retained to the present day. Currently, it is operated by the Japanese Society of School Health. As of the end of 2019, the system covered approximately 30,000 schools, which collectively account for about 60% of all schools in Japan. It also covers approximately 10,000 nursery schools, which collectively account for about one-third of all nursery schools nationwide. Every day, it monitors the health condition of about 6 million people younger than 18 years old.

The present study has some limitations. The first is that because our case definition used for this study was based on clinical diagnosis and because it was not based on test results, misdiagnoses for patients with herpes zoster or other (infectious) diseases such as invasive group A streptococcal infection [18] or hand–foot–mouth disease might be included among the varicella patients.

A second limitation was that after one child was diagnosed as being infected by varicella, staff members at nursery schools search intensively for a rash on the children’s skin. By that process, they found very mildly infected patients with very mild symptoms, e.g. with fewer than five rashes. Conversely, outside of a nursery school, children’s skin was not checked so intensively. Therefore, some might have otherwise been found to be varicella patients. At nursery schools, the perceived and recorded incidence might be higher. If that were true, then VE might be lower than that of children who did not attend nursery school, in addition to their differences in contact.

A third limitation is related to immunization records. In principle, the data are based on information recorded in maternity passbooks, as described above. However, when caregivers register immunization records, they might misunderstand information included in the maternity passbook. If so, then the variance in VE can be expected to be large. It would therefore become difficult to obtain significant results.

## Data Availability

We do not have any data available in public in the present study.

## Acknowledgments

We acknowledge all participants at nursery schools at studied area. This study was supported in part by a grant from the graduate school of Tokyo Kasei University and Tokyo Women’s Medical University.

